# Multilevel Integrated Model with a Novel Systems Approach (MIMANSA) for Simulating the Spread of COVID-19

**DOI:** 10.1101/2020.05.12.20099291

**Authors:** Arpita Welling, Abhilasha Patel, Padmaj Kulkarni, Vinay G. Vaidya

## Abstract

Due to the spread of the coronavirus, public health officials grapple with multiple issues such as recommending a lockdown, contact tracing, promoting the use of masks, social distancing, frequent handwashing, as well as quarantining. It is even more challenging to find the optimal combination of these factors without the use of a suitable mathematical model.

In this paper, we discuss a novel systems approach to building a model for simulating the spread of COVID-19. The model, MIMANSA, divides an individual’s in-person social interactions into three areas, namely home, workplace, and public places. The model tracks the in-person interactions and follows the virus spread. When a new silent carrier is created, the model automatically expands and builds a new layer in the network.

MIMANSA has four control mechanisms, namely the exposure, infection rate, lockdown, and quarantining. MIMANSA differentiates between virus-infected patients, silent carriers, and healthy carriers. It can consider variations in virus activity levels of asymptomatic patients, varying the exposure to the virus, and varying the infection rate depending on the person’s immunity. MIMANSA can simulate scenarios to study the impact of many different conditions simultaneously. It could assist public health officials in complex decision making, enable scientists in projecting the SARS-CoV-2 virus spread and aid hospital administrators in the management of beds and equipment.

MIMANSA is trained and validated using the data from the USA and India. Our results show that MIMANSA forecasts the number of COVID-19 cases in the USA, and India within a 3% margin of error.

## 1 Introduction

Mathematical modeling of COVID-19 (Coronavirus Disease 2019) is essential for many reasons. It helps public health officials understand how COVID-19 may grow, predict the number of cases in a given region, and prepare better for patient management. Another important aspect is that a good model can enable administrators to implement the right public policies and respond to changes in real-time using data-driven models. The models can provide assistance in answering questions related to the effect of an increase in the number of cases, the impact of lockdown, lifting of the lockdown, mask usage, social distancing, contact tracing, and isolation.

There are many attempts made to develop mathematical models of COVID-19; however, they do not always answer questions related to the impact of multiple measures and policies.

In this paper, we present a novel approach by considering the SARS-CoV-2 (Severe Acute Respiratory Syndrome Coronavirus 2) infection as a conglomeration of multiple systems. Our model, Multilevel Integrated Model with A Novel Systems Approach (MIMANSA), is built from epidemiological observations and data. It is well known that studying the asymptomatic population is vital in the study of the spread of COVID-19. The simulation of the spread through silent carriers is at the heart of MIMANSA. Inside the model, MIMANSA can delineate between patients infected with the virus as well as those that are asymptomatic or mildly symptomatic.

The model distinguishes the degree of spread based on an individual’s interaction between three cohort groups, namely individuals at home, workplace, and public places. It is built on globally acceptable observations related to the spread of SARS-CoV-2. It has provisions for simulating non-pharmaceutical interventions such as a lockdown or quarantine. It can also simulate scenarios to study the impact of following precautions such as mask usage, social distancing, and frequent hand washing. Once trained, it can also highlight the deviations of lockdown compared to the nominal. It has multiple features that enable studying several different scenarios.

## 2 Literature Survey

There is a vast amount of literature available on the curve-based models. These models include exponential, Gompertz, Bertalanffy, and Logistic. All of these have been tried for COVID-19 simulation.

The exponential model is one of the most widely used models for the growth of any organism. Although suitable for measuring early behavior, the model starts deviating rapidly as time progresses. The growth in COVID-19 patients does not follow the exponential curve. Its nature is more like that of a 3rd order polynomial. Villalobos and Mario [1], use a generalized logistic equation and the Gompertz equation for fitting COVID-19 data from China.

Jia et al. [2] use Gompertz, Bertalanffy, and Logistic equations for modeling the growth of COVID-19; however, they do not help in considering multiple scenarios that arise in the management of COVID-19 such as the lockdown, quarantine, exposure variation, etc.

Time series analysis, using the Auto Regressive Moving Average (ARMA) models, has been another popular approach. This approach has been tried by Deb et al. [3] on COVID-19. Although it gives initial success in prediction, the success does not last long when the growth rate of cases suddenly changes due to either a sudden outbreak or due to a group of people getting together in large numbers. There is no provision for adding new clusters of silent carriers in the ARMA model. If there is a sudden increase in the numbers due to unknown external factors, one has to recalculate the entire model.

Mandal et al. [4] used the classical SEIR (Susceptible, Exposed, Infectious, and Recovered), model. They suggested that screening at the port of entry for India may result in some delay in getting the coronavirus in the country. However, screening at the port alone may not suffice in delaying the outbreak. Fanelli and Piazza [5] forecasted cases in China, Italy, and France based on the variation of the SEIR to the SIRD model. In this approach, the Susceptible (S), Infected (I), Recovered (R), and the Dead (D) model, every person who is going to be infected by the virus, falls in one of the four categories. Then the equations are set up as first-order differential equations with one differential equation per stage.

Wang et al. [6] present their work on the transmission dynamics of SARS-CoV-2. They used the SEIRD (Susceptible, Exposed, Infectious, Recovered, and Dead) model for the Wuhan data.

Kucharski et al. [7] studied the early dynamics of transmission. They modeled SARS-CoV-2 using a geometric random walk process and Monte Carlo simulation. Grassly and Fraser [8] have given a good review of mathematical models of infectious disease transmission. Klinkenberg et al. [9] developed a model for contact tracing.

The systems approach was first suggested by Von Bertalanffy [10]. In this approach, a system is broken down into small subsystems. The subsystems are open to interacting with the environment as well as with each other. One time you may see parts of the whole system while other times, you may like to view the whole system from parts. Each subsystem is built on observed facts. All subsystems integrate together and keep the observed phenomenon unperturbed. The systems approach has been effectively used for developing a model for tumor growth and chemotherapy and is given in [11]. Later, in 1991, the effect of fasting on reducing tumor growth by using a systems approach was shown in [12]. The ‘Multilevel Integrated Model with a Novel Systems Approach’ (MIMANSA) is built using the system’s principles.

## 3 Mathematical Model

The MIMANSA model developed here is based on empirical data and global epidemiological observations of the COVID-19 cases. The observations used while building the model to explain the primary virus spread are given below. The virus spread occurs due to coming in contact with a virus-infected or virus-carrying person.

- The silent and healthy carriers are responsible for the spread of the virus.
- A healthy person may get infected if the person touches a fomite.
- For all reporting, the unit of time is one day. The total number of patients is reported every day. Similarly, new silent carriers are created every day.
- As there is a 2 to 14-day incubation period for the SARS-CoV-2 virus, virus-infected persons do not show symptoms on day 1. It is observed that most patients start showing symptoms on the 5th, 6th, or 7th day after being infected, with 97.5% showing symptoms by 11.5 days. [13, 14].

The following assumptions are made for the development of the model.

1. Infected patients spread the virus only during the incubation period while they are asymptomatic. When symptoms show, it is presumed that the patients will be hospitalized or effectively isolated/quarantined, eliminating them as a vector that can spread the virus to other healthy individuals. Silent carriers and healthy carriers with fomite are the ones who spread the virus.
2. The SARS-CoV-2 virus remains in aerosols for up to 3 hours, copper surfaces for up to 4 hours, and on cardboard for 24 hours. On plastic and stainless steel, it can remain for up to 2 to 3 days [15]. For simplicity, we consider the period of the virus remaining on a surface to be 24 hours.

To understand the spread of diseases, Del Valle et al. studied how people from different age groups interact socially [16]. In their study, they found that on average, between the ages of 20 and 50, a person meets 22 people every day. We have used this number to simulate the spread of SARS-CoV-2.‘

In Fig. 1(a), a person P meets a silent carrier P1 and is infected with the virus. P interacts in three groups, namely family, workplace, and public places. It is important to note that there is only one individual, i.e. P in the example, who interacts with people in all three groups. In all, P meets 22 people. All events are assumed to have taken place in one day, the time unit for the simulation cycle.

**Fig. 1(a):**
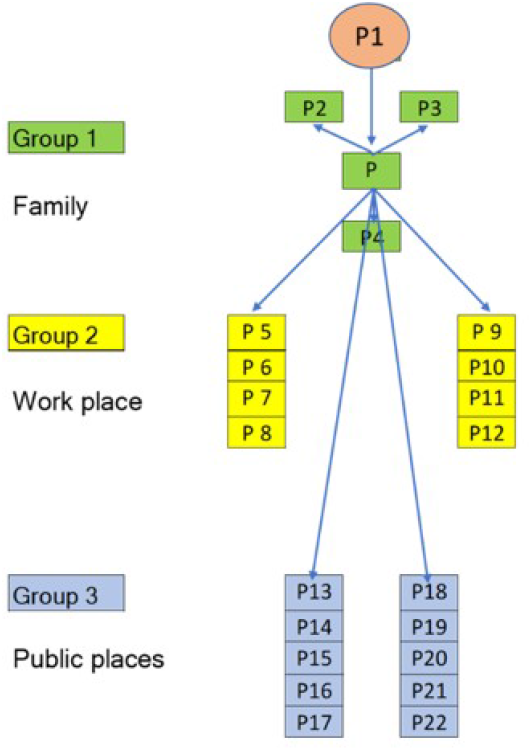
A person P meets 22 people in a day

### 3.1 Definitions

Some of the important terminologies used in this paper are given below. The remaining acronyms are given in Table 1 and Table 2.

**Table 1:** Definition of the terminology used.

| Terminology Used | Definitions with explanation |
| --- | --- |
| CB | Computational block inside which calculations for $(L, t)$ take place. |
| CPDFVP | Cumulative Virus Patients after passing through incubation emulating PDF |
| CTQ | Contact Tracing, and Quarantining parameter varies from 0 to 1. |
| CVP | Cumulative Virus-infected Patients is termed as CVP |
| ER | The exposure rate is the probability of getting exposed while interacting with silent carriers. The ER (1), ER (2), ER (3) are exposures for 3 groups. |
| $F(L, t)$ | Fraction of virus patients with symptoms to the total number of virus patients. |
| G | In-person, social interaction Group |
| HC1 | Healthy Carriers, carrying fomites, coming from group 1 |
| HCR | Healthy Carrier Rate. An infected person due to coming in contact with a fomite is called the Healthy Carrier Rate (HCR). |
| HVP | Hidden Virus Patient |
| Input 1 | The number of silent carriers at the start of the simulation |
| IPD | Inter-Process Diagram outlines internal calculations |
| IR | IR is the probability of getting infected if one is exposed to the virus. |
| L | Levels indicate the depth of the infection network |
| LKD (LKD2, LKD3) | LKD is the fraction of the people adhering to the lockdown norms. The suffix indicates a group. |
| $N_1, N_2, N_3$ | The number of people involved in interactions in every group is called N. |
| PDF | Probability Distribution Function |
| PDFSC | Silent Carriers coming out PDF |
| PI | People Infected and presently carriers |
| PSCP | Probability-based Silent Carriers ready to Proliferate on the next day |

**Table 2:** Definition of the terminology used.

| Terminology Used | Definitions with explanation |
| --- | --- |
| SCE | External Silent Carriers |
| SCP | Silent Carriers at Present for starting their virus spread network |
| SCR | Silent Carrier conversion Rate. |
| SHVP | The Sum of the Hidden Virus-infected Patients in the last 14 days |
| SSC | The Sum of the Silent Carriers is SSC |
| t, T | The time, in days, from the start is t. T is the incubation cycle time. |
| TCTQ, TSC | TSC, Total Silent Carriers. TCTQ is the enforcement time of CTQ. |
| VP, VP1, VP2, VP3 | A Virus-infected Person tested positive for COVID-19. The suffix attached to any variable indicates the group from where the infection occurred. |
| VPPDF | Virus-infected people coming out of the PDF |

A Virus-infected Person (VP) is a patient that is evaluated clinically and tested positive for COVID-19.

A Silent Carrier (SC) is a person who is infected with the virus but is not considered a patient. The silent carriers include persons who are asymptomatic carriers and those who show mild symptoms that do not get tested.

A Healthy Carrier (HC) is a person who is not infected with the virus but carries the virus via fomites. Fomites are materials such as a book, a bag, clothes, utensils, etc. that have been contaminated by the virus and serve as a mode of transmission.

For the three groups, we define three levels of Exposure Rates (ER). The ER (1) is the highest exposure level between members of an individual’s family. The ER (2) is that between the individual and office members, and ER (3), the lowest exposure level experienced between the individual and members of the general public in congregate settings. The exposure rate is the probability of getting exposed while interacting with silent carriers.

In table A, we have given a detailed list of the terminology and definitions used in this paper.

### 3.2 Internal Processing Diagram (IPD)

Fig.2a is an Internal Processing Diagram (IPD). This diagram shows what happens to every healthy person who comes in as a part of the group interactions. Inside the Internal Processing Diagram, the input is converted into three outputs, namely Virus-infected Patients (VP), Silent Carriers (SC), and Healthy Carriers (HC).

**Figure 2a:**
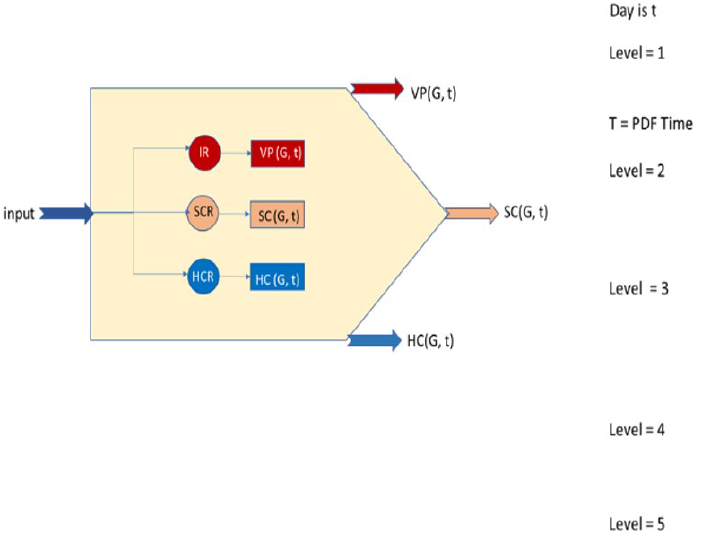
Internal Processing Diagram - the division of people as per their state of infection

The virus-infected patients have an incubation period of 14 days and may show symptoms anytime within the 14-day window [17]. In MIMANSA, we use a probability density function to simulate the same effect as the incubation period.

Based on initial epidemiological data from the WHO QA session on March 17, 2020, 80% of infections are mild or asymptomatic, 15% develop severe symptoms requiring hospitalization, and 5% deemed critical requiring artificial ventilation. It is also reported that 50% of the patients are asymptomatic [18]. Using this data, we were able to define broad ranges for Infection Rate (IR), Silent Carrier Rate (SCR), and Healthy Carrier Rate (HCR).

### 3.3 Computational Block

Fig. 1b shows the internals of a Computational Block (CB). All calculations in a computational block are equivalent to the activities taking place in a single day.

**Fig. 1(b):**
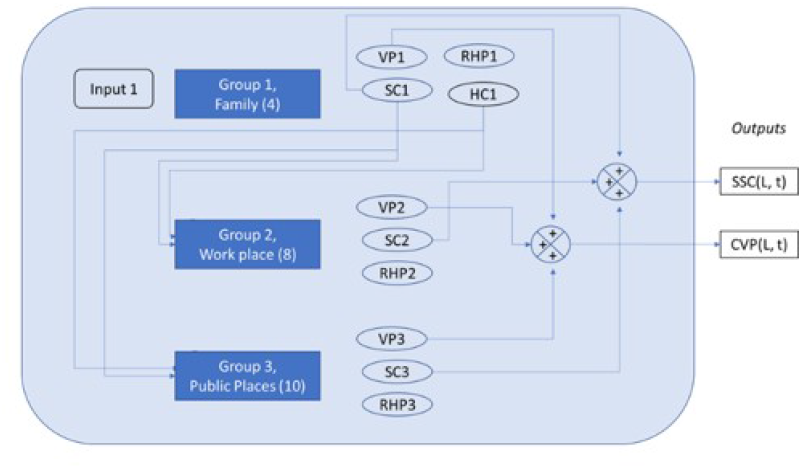
The people met are arranged inside a CB, the Computational Block

Inside a computational block, there are computations for each of the inperson interactions within each group 1, 2, and 3. The output of a computational block is the total number of silent carriers (SSC) and Virus-infected patients (CVP). The healthy carriers are not carried forward to the next day cycle since we have presumed that a fomite can infect only for a 24-hour duration.

All computational blocks are integrated at the end to get the final result. The only interaction between the blocks is to transmit the virus through the silent carriers who meet yet another group of family, colleagues, and individuals in public places.

### 3.4 The Virus Proliferation Network

When one day is over, the Sum of the Silent Carriers (SSC), and Cumulative Virus-infected Patients (CVP) are reported.

The next day, the person P, in our example, meets the same people from group 1, and group 2. It is likely that P meets different people from public areas. However, for simplicity, it is assumed that P meets the same people from the group 3 as well if one goes to the same public places such as grocery store, bus or a train station, and meet the same people. From each of these three groups, some people get infected on day 1 itself, but not all of them do. The remaining healthy people from the previous day, again interact the next day with the same silent carrier. Thus, we need to treat these remaining healthy people differently. In our simulation, we transfer the remaining healthy people to another horizontal block, and that is the day 2. In a horizontal block shown in Fig. 2b, we have the same people interacting with each other.

**Figure 2b:**
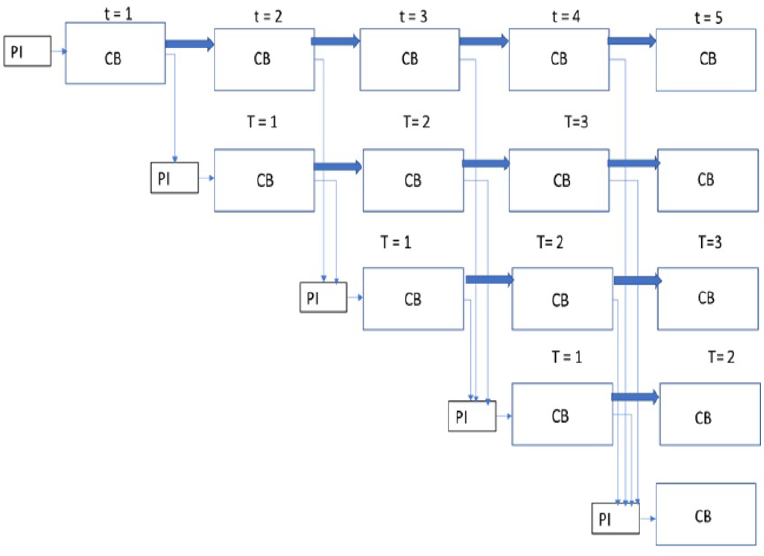
Virus Proliferation Network Using the Computational Blocks, one for each day and level.

Each day, the chain of spread from the newly created silent carriers, created from the previous day prior, is represented on a new level. Consider, for example, P6 from group 2, and P12 from group 3 have become silent carriers on day 1 (Fig. 1 a). Both P6 and P12 have their respective families and a different network of people they interact, averaging 22 interactions each. To represent that, we create a new level, level 2. This is a vertical block in Fig. 2b. Vertical blocks are created whenever there are new possibilities for the in-person interactions in the network. The new levels are started by all the silent carriers coming from the top levels. If we are at level 20, we add all the silent carriers from level 19 and going back to level 5. This covers 14 levels in the past. The reason for going all the way up to 14 levels and no more is due to the incubation period of SARS-CoV-2 being 14 days. There is no silent carrier that would continue to be a silent carrier beyond 14 days. This is how the virus spreads its network. In a horizontal block, it is the intra-group transmission while in a vertical block, it is the inter-group transmission.

### 3.5 Virus Spread within the family

In MIMANSA, the process of virus spread starts with the first silent carrier who comes from abroad (Fig. 1a). Input1 in Fig. 3a describes the arrival of the first silent carrier. In the simulation, there is a provision for multiple silent carriers coming into a place and starting the process. The input 1 is multiplied by the first group of healthy people (N1), that the silent carriers meet. These people pass through an IPD. Once exposed to silent carriers, healthy people are marked in the IPD as Virus-infected Patients, new Silent Carriers, or Healthy Carriers. Every IPD symbol has these three outputs. Some healthy people remain healthy despite getting exposed to a silent carrier, and they are marked as the Remaining Healthy People. Throughout the paper, color red indicates a virus-infected person –related box, blue indicates healthy people, and brown indicates a silent carrier-related box.

**Figure 3a:**
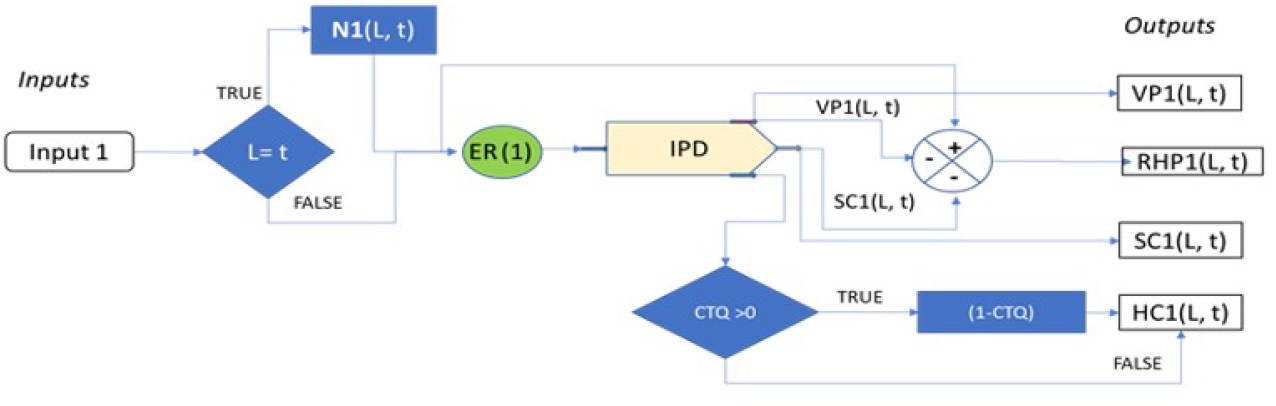
Group 1: The virus spread within the first group

### 3.6 Virus Spread at Workplaces

The next figure, Fig. 3b describes the spread of the virus in group 2, i.e., workplace.

**Figure 3b:**
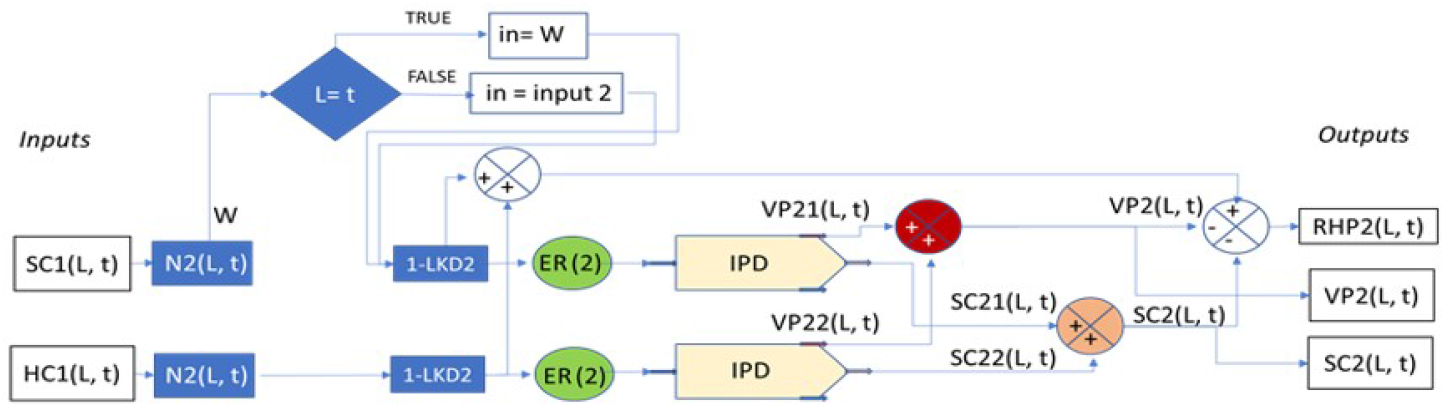
Group 2: The virus spread within the second group

Group 2 is designated as the workplace group. The virus spreads at a work-place due to the in-person interactions with a silent carrier. The inputs to the group 2 come from silent carriers and healthy carriers from group 1 if the computations are in a horizontal block. The horizontal block means that the same silent carrier is interacting among the same people they interacted the day before. W is an intermediate variable whose value is the product of SC1 and N2.

In Fig. 3b, the concept of lockdown is introduced. If the lockdown parameter (LKD2) is 0.6, it means that the lockdown was 60% adhered to. This lockdown, LKD2, is applicable only to the workplace group. Thus, if a company has a work from home policy, and everyone follows it, the LKD2 can be set to 1, i.e., 100% lockdown. Also, there is a division in computations of Virus-infected Patients (VP) and Silent Carriers (SC) as VP21 and VP22, and SC21 and SC22. The virus spread is split into two categories. Category 1 indicates that the virus is spreading due to the Silent Carriers (SC). Category 2 means that the virus is spreading due to Healthy Carriers (HC). Thus, SC21 means silent carriers created in group 2 with category 1, and SC22 means that the silent carriers are created in group 2 with category 2.

Outputs of this group 2 are Virus-infected Patients (VP), Silent Carriers (SC), and Remaining Healthy people (RHP). They are used for further processing. This is explained in subsequent diagrams.

### 3.7 Virus Spread in Public Places

In Fig. 3c, we describe the virus spread in public places. The group3 is marked as the group of people one meets at public places. The only difference between the group2 and group 3 is in the values of ER. There are many commonalities between the virus spread diagram for group 2 and group 3. The main distinction is in the exposure rate used for healthy carriers spreading the virus. Since the group 3 is the interaction in public places, the duration and the level of contact is different. Thus, we have used ER (3) as the exposure rate. Outputs from group 3 are used in subsequent computations. W is an intermediate variable whose value is the product of SC1 and N3.

**Figure 3c:**
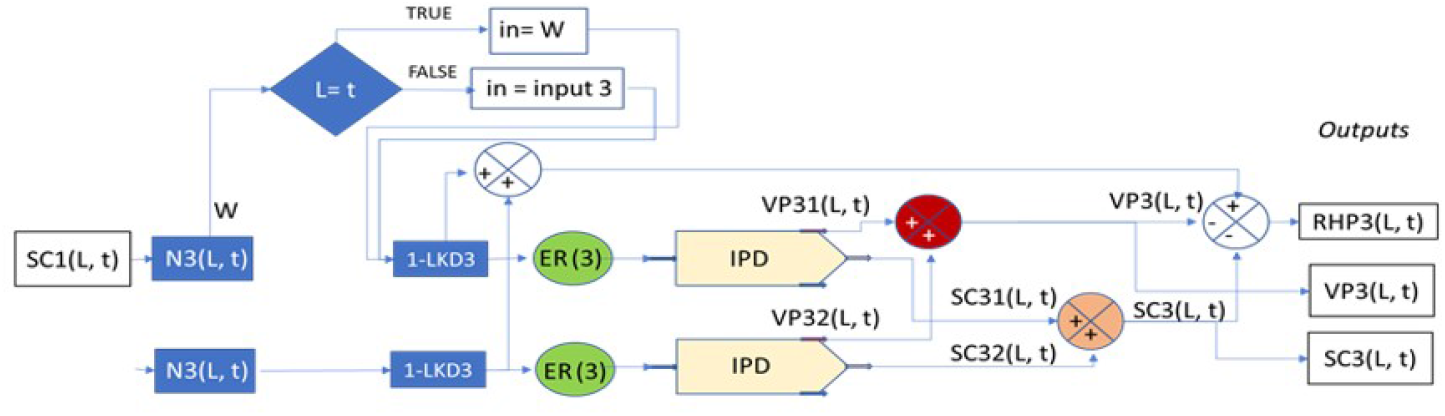
Group 3: The virus spread within the third group

**Figure 3d:**
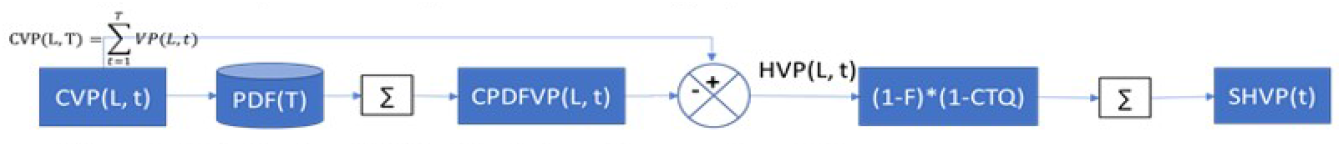
Distribution of Hidden Virus-infected Persons using a PDF

ER (2) is in an office environment. The number of people per square feet is going to be less than going on a train, bus, or a shopping center. Thus, ER (3) will change according to the type of place one meets. It is assumed that ER (2) ¿ ER (3) due to the amount of time spent.

### 3.8 The Equations

Mathematical equations for VP, SSC, RHP when all the three groups are put together, is shown below.

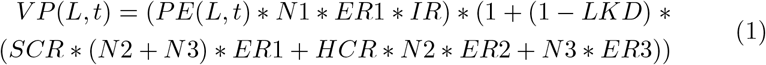

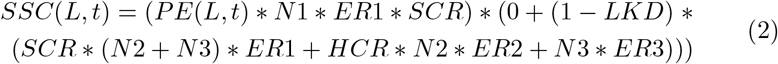

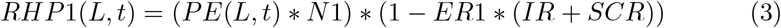

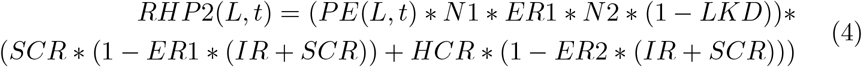

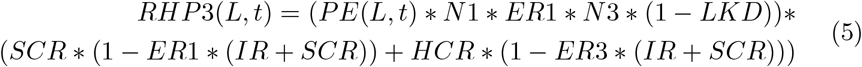

These equations get a bit more complex once a probability distribution functions are applied on VP (L, t) and SSC (L, t). How to apply a probability distribution function is outlined next.

### 3.9 Simulating the Incubation Period by a PDF

In the computational block, CB, shown in Fig. 1b, we have shown VP (L, t) as one of the outputs. However, all virus-infected persons do not show up in one day. Thus, we have used a probability distribution function PDF(T) that operates on VP (L, t). A person exposed to the virus may exhibit a high rate of infection at any time between 2 to 14 days. It is observed that 5 to 7 days is the estimated mean incubation period [14]. To emulate the incubation period of SARS-CoV-2, we designed a PDF in line with the observations. This makes it possible to simulate patients showing symptoms slowly.

The concept of the incubation period is not only applicable to people identified to become virus-infected persons, but it is relevant to silent carriers as well. Thus, we designed another PDF for silent carriers.

The sum of total virus patients from each of the three groups in a Computational Block is called Cumulative Virus Patients (CVP). Each Computational Block gives us a different CVP count. After we distribute the CVPs through PDF, we get the total number of virus patients at time t and level L. The total number of virus patients, is called the Cumulative of PDF output as a Virus-infected patient, CPDFVP.

From the output of the Internal Processing Diagram, some people are marked as the ones who are going to be Virus-infected Patients. We repeat that these individuals are only marked internally. They have the infection, and they are silent carriers. When the incubation period is over, they start showing symptoms and go to a hospital. However, until they come out as Virus-infected Patients, they are not known as patients, or they are hidden. These patients who are not yet showing symptoms are the Hidden virus patients (HVP). HVPs for a certain Computational Block (CB) can be found out by subtracting Cumulative PDF output of Virus-infected Patients CPDFVP (L, t, i) from the Cumulative Virus-infected patients, CVP (L, t) of that Computational Block (CB). The first block of every new level gets HVP as one of its inputs. Since the incubation period is 14 days, we give the Hidden virus patients (HVP) as input only for the last 14 levels. For example, a new Computational Block (CB) with time 16 and level 16, will get HVPs from CBs at time 15 and levels 2 to 15. The sum of 14 latest HVPs is called Sum of Hidden Virus Patients (SHVP) as in equation 6.

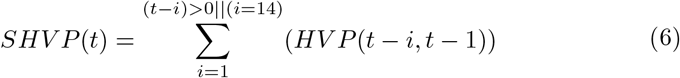

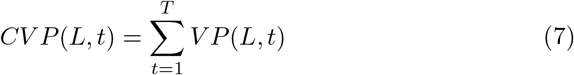

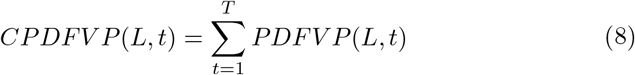

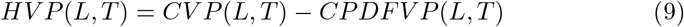

The silent carriers of group 2 and group 3 of CB are together called as Sum of Silent Carriers (SSC).

There are different opinions about the role of asymptomatic patients in the spread of SARS-CoV-2. The initial thought process was that asymptomatic patients do not spread the virus. However, now there is evidence that asymptomatic people can infect others (Bai et al., [19]) (Hu et al. [20]). Since there is still no concurrence on the percentages, we consider that the virus transmission from symptomatic patients is 20%. We have also considered that asymptomatic patients are 50% of all the patients. Thus, 20% of 50% gives 10% of the total. This is the reduction factor (RF). Further, if we consider the case of family quarantine, we also multiply SSC by the quarantine factor (1-Q), where is Q is the Quarantine flag. The quarantine flag is 0 if there is no quarantine. If the quarantine policy is applied, then the quarantine flag is equal to the Contact Tracing and Quarantine value (CTQ). Thus, we get Total Silent Carriers (TSC) when we apply reduction factor (RF) and 1-Q to Sum of Silent Carriers (SSC). The first block of every new level gets TSC as one of its inputs. But we only give the TSCs of the last 14 levels as input. For example, a new CB with time 17 and level 17 will get TSCs from CBs at time 16 and levels 3 to 16. The sum of 14 latest TSCs is called Silent Carrier at Present (SCP).

The formula is given by:

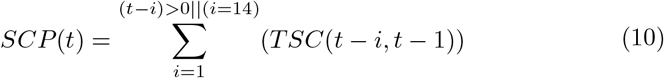

where SCP = Silent Carrier Population, t-i = Level of computation at time t, TSC = New or incremental Silent cases, t = Current day count.

As all Sum of Hidden Virus-infected Patient (SHVPs) and Silent Carrier at Present (SCPs) do not start infecting people at the same time, we distribute them over 14 days using a Probability Distribution Function (PDF), as shown in figure PDF Graph. The distributed SHVP and SCP are together called as Probability Distribution based silent carriers and hidden virus patients (PDFSC). At every new level, we take the total Sum of Hidden Virus-infected Patient (SHVP) and Silent Carrier at Present (SCP) that are active at that time and together call them PSCP.

As shown in Fig. 3e, PSCP, along with external silent carriers (SCE), form the People Infected (PI), which serves as the input for the first block of every level. For the further blocks of a level, we first check if a patient has already been detected and if family quarantine is implemented. If that is the case, then our input is cut off, and that level will not execute further. But if a virus patient is detected but family quarantine is not implemented, or vice-versa, we give the 3 inputs to CB as RHP1, RHP2, RHP3 from the previous blocks.

**Figure 3e:**
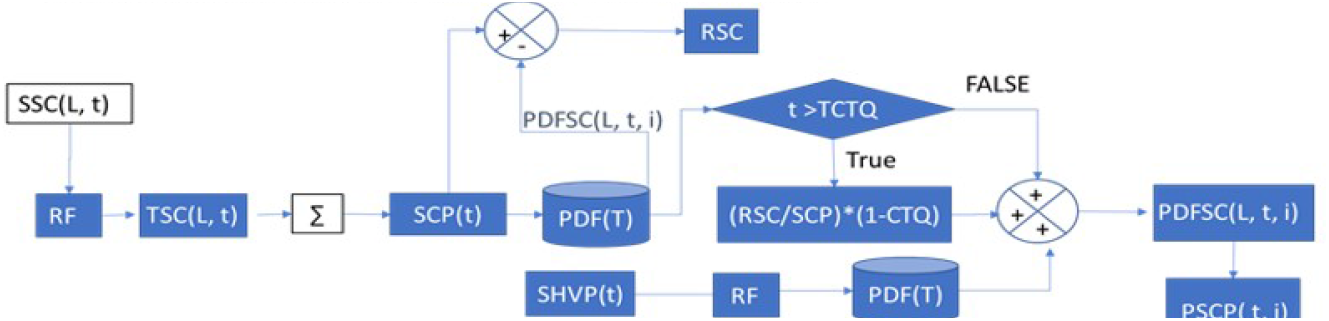
Distribution of Silent Carriers using a PDF

In the next figure, Fig. 3f, input calculations are described. The cycle of virus spread continues from one day to the next. The computational block, shown in Fig. 1b represents the simulation of a single day. The next day, a new iteration starts, and new calculations are made. In terms of the virus spread, this means that the next day the virus continues to spread. The spread during the next day depends on the count at the end of the previous day. Virus growth is a self-propelling system.

**Figure 3f:**
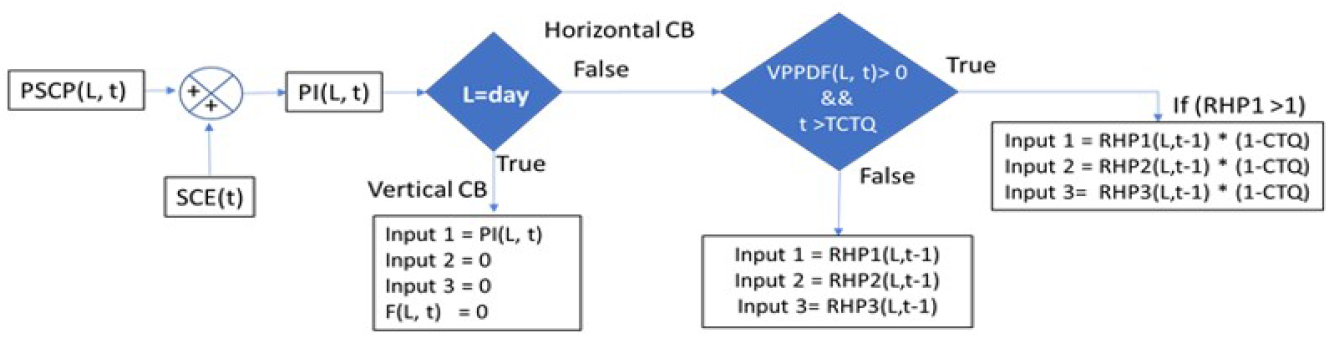
Input calculations for the next iteration

Since we are building a self-propelling system, it has to have internal inputs that keep feeding back on the next day. In Fig. 3f, we show how these new inputs are calculated based on the old outputs.

### 3.10 Inputs for the Next Cycle

Fig. 3 f shows the different possibilities of inputs. The inputs may be to a horizontal block or a vertical block shown in Fig. 2b and explained in the section Virus Proliferation Network. Remaining healthy people are passed from one block to another horizontal block. In case there is a contact tracing and quarantining policy, it can be controlled through the variable CTQ. If a new vertical block is started, its input is PI (L, t).

## 4 Methods and Materials

In this study, we have developed a model to study the number of cases of COVID-19. The total system diagram involves multiple parameters. We have derived some parameters based on actual data available on the COVID19India.org and Kaggle.com website [21], while the values of other parameters are taken from literature. In some cases, we had to use the best judgment to ensure that the values are likely to occur in real life. The following is a list of assumptions.

1. The data used for training the India model was from March 1, 2020, up to May 25, 2020, while the USA was from January 22, 2020, to May 25, 2020. Validation in both cases was done until June 5, 2020.
2. All workplaces, except for essential services, were closed during the lock-down in India. The terminology lockdown is same as the stay at home order in the USA. This lockdown started from March 25, 2020; however, slowly, people started abiding by it. Thus, we have gradually changed the value of the lockdown percentage. However, in the case of the USA, lockdowns started at different times, depending upon the state. So, in our simulation for predicting nationwide patient count, we have assumed dates by considering the population, population density, number of lockdowns occurring on that date, number of lockdowns ending on that day, etc.
3. The Infection Rate (IR) was calculated using the data from COVID19India [21]. Extensive contact tracing was done in India. All data related to the traced contact is also part of the dataset. Thus, one can find out how many people were infected by one individual. Using Del Valle’s study [16], we know that, on average, a person meets 22 people. By taking the ratio of people infected to the number of people met, one gets IR. The calculated value of IR is 0.12.
4. The number of people in each of the 3 groups are selected as follows. In general, it is reasonable to assume that a family consists of 4 people. At the workplace, one meets a limited number of people compared to in public places. As per Del Valle [16] one meets 22 people every day. With that total in mind, we select 4 people at home, 8 people at the workplace, and 10 people in public places. Although these numbers may vary in individual cases, on average, it appears to be a reasonable distribution.
5. For selecting the exposure rate, we calculated the actual exposure rate among Italian doctors. In a survey, (8), Italian doctors reported that 108 out of 272 had symptoms of COVID-19. Out of the 272 GPs, only 125 had direct contact with a confirmed COVID-19 patient. The ratio of those who had symptoms to those who were exposed is 0.864. Since ER (1) indicates the highest level of exposure, we can choose a number close to 0.78.
6. All exposure rates and the silent carrier rate were modeled as a function of the lockdown. The equations used were:
  a. ER (1) = m1*Lockdown + c1
  b. ER (2) = m2*Lockdown + c2
  c. ER (3) = m3*Lockdown + c3
  d. SCR = m4*Lockdown + c4
7. The Healthy Carrier Rate (HCR) is taken low since the possibility of fomite transmitted infection is low.
8. The lockdown values were based on the news reports.
9. We consider 80% silent carriers, 15% hospitalized, and 5% needing ICU care. This is based on the study by Wu and Mc Googan [22] and as per the WHO QA session [23] on March 17, 2020.
10. Asymptomatic patients can transmit the virus, even if they continue to show negative on the SARS-CoV-2 test. [24].
11. We conducted several simulations to get the best fit for the USA and India data. Every parameter was adjusted carefully by keeping the values within the practical limits to ensure that the best-fit parameters are not merely a numerical play, but represent reality as closely as possible.

## 5 Results and Analysis

The following results and discussion are about the COVID-19 cases in the USA and in India. The actual data was obtained from the Kaggle website[25] for the USA and COVID19India.org website for India[21].

Figures 4 and 5 show the training, predictions, and lockdown variations in India and the USA. Please note that the terms ‘lockdown percentage’ and the ‘change in mobility in percentage’ are synonymous. The parameter values, after the best fit, are given in Appendix A, Table 3. The start and end dates for the training data for the USA were January 22, 2020, to May 25, 2020, and for India, they were March 1, 2020, to May 25, 2020. As can be seen in the figures below, MIMANSA values match reasonably well to the actual data. For both India and the USA, predictions up to August 15, 2020, are shown in the figures. These projections are based on several assumptions, including lockdown variations, availability of vaccines, new findings of the virus spread, etc. Thus, the predictions are indicative and not necessarily accurate.

**Fig. 4:**
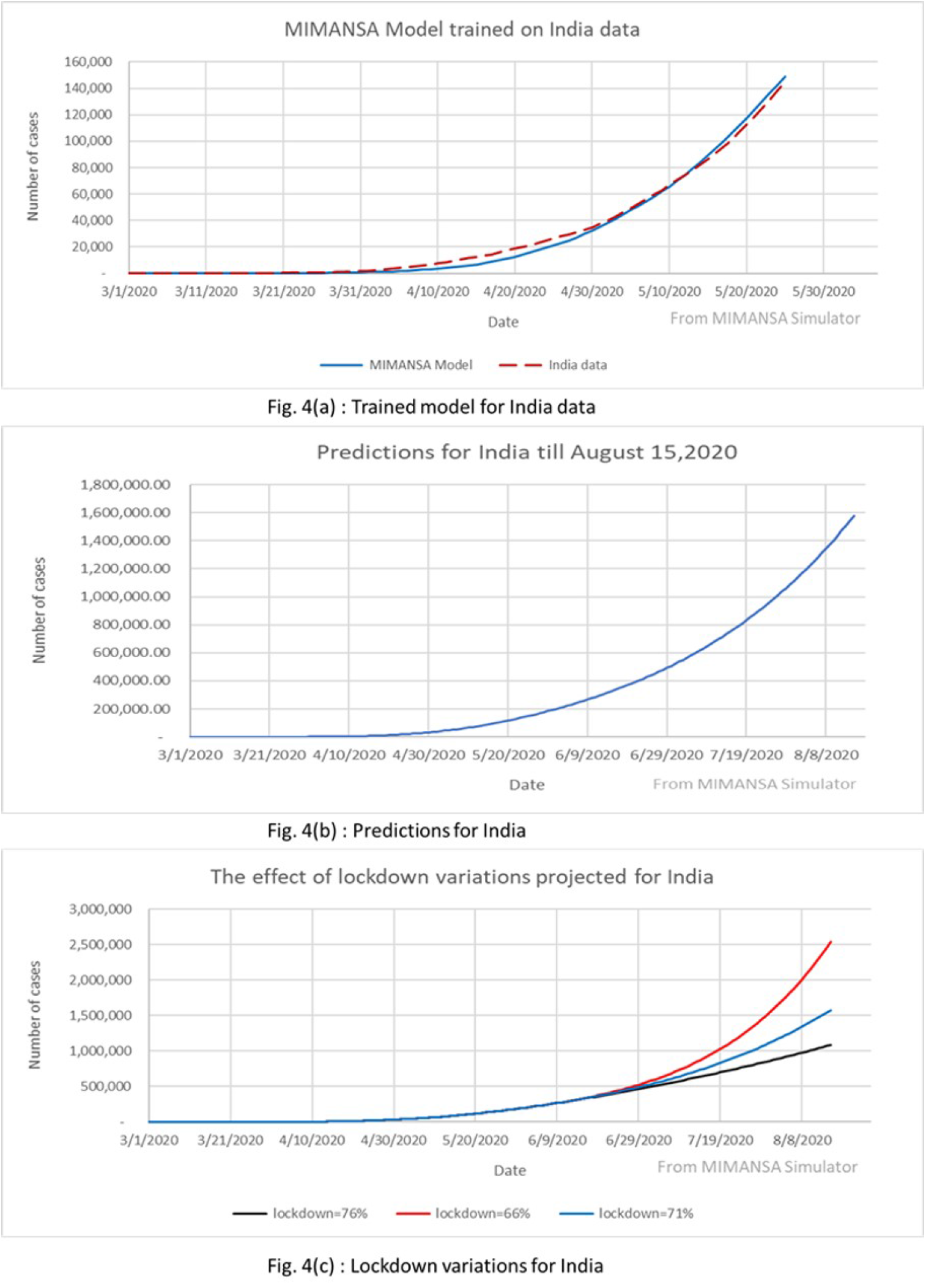
Training, Predictions, Lockdown variations for India

**Fig. 5.**
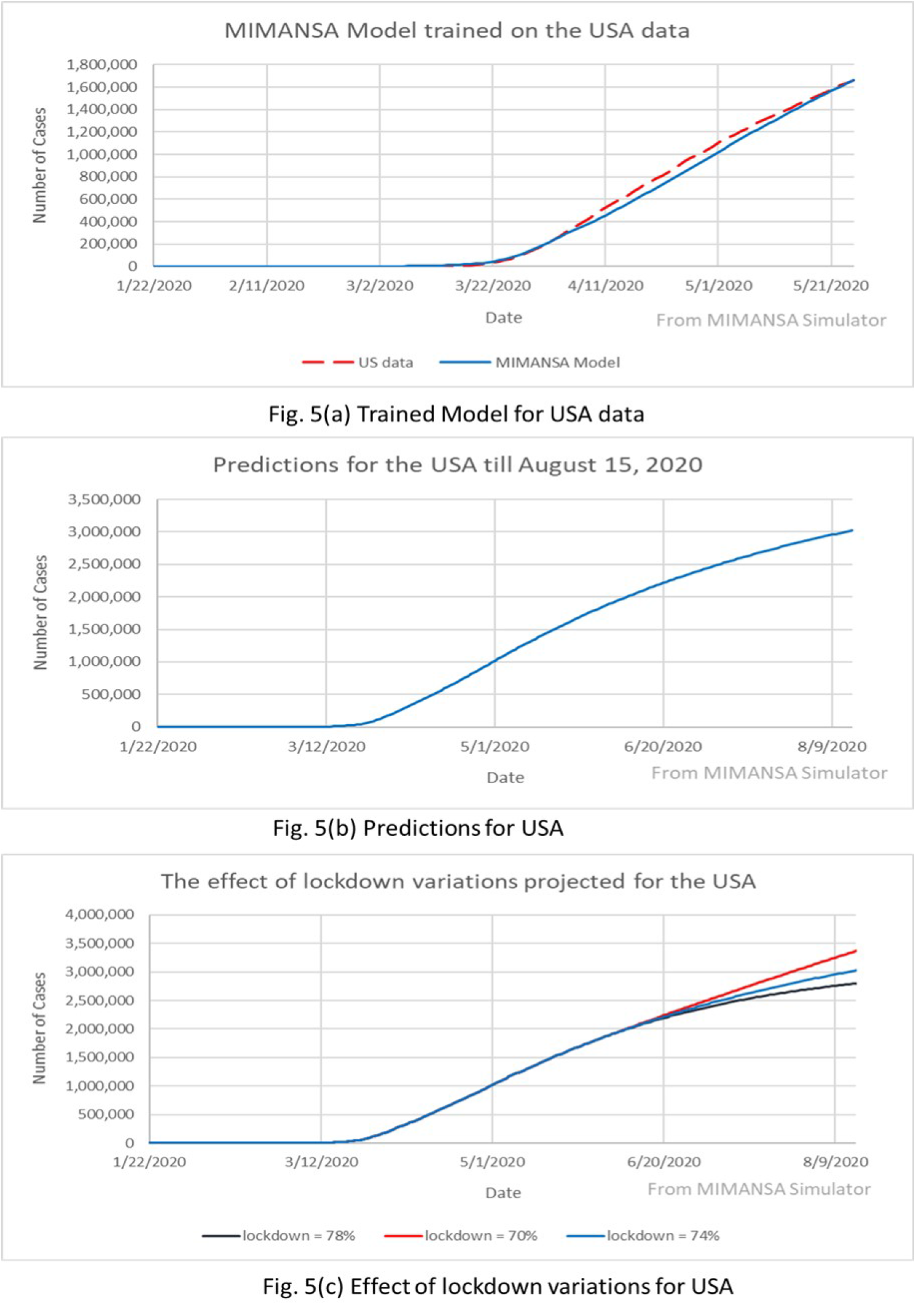
Training, Prediction and lockdown variations for the USA

**Table 3:** The table shows all the model parameters for US and India data.

| Parameter | Values for the USA | Values for India |
| --- | --- | --- |
| N1 | 4 | 4 |
| N2 | 10 | 10 |
| N3 | 8 | 8 |
| IR | 0.12 | 0.12 |
| HCR | 0.12 | 0.12 |
| RF | 0.1 | 0.1 |
| m1, m2, m3, m4 | -0.1484, -0.1659, -0.1659, -0.1659 | -0.1484, -0.1659, -0.1659, -0.1659 |
| c1, c2, c3, c4 | 0.56484, 0.46369, 0.41143, 0.54304 | 0.75, 0.603, 0.5037, 0.536 |
| First Lockdown Start Date | 19-Mar-20 | 16-Mar-20 |
| Second Lockdown Start Date | 23-Mar-20 | 25-Mar-20 |
| Third Lockdown Start Date | 30-Mar-20 | 4-Apr-20 |
| Fourth Lockdown Start Date | 3-Apr-20 | 17-Apr-20 |
| Fifth Lockdown Start Date | 30-Apr-20 | 29-Apr-20 |
| Sixth Lockdown Start Date | 8-May-20 | 17-May-20 |
| First lockdown (LKD 2, LKD 3) | 0.1 | 0.1 |
| Second lockdown (LKD 2, LKD 3) | 0.28 | 0.3 |
| Third lockdown (LKD 2, LKD 3) | 0.53 | 0.37 |
| Fourth lockdown (LKD 2, LKD 3) | 0.66 | 0.55 |
| Fifth lockdown (LKD 2, LKD 3) | 0.75 | 0.63 |
| Fourth lockdown (LKD 2, LKD 3) | 0.74 | 0.71 |

Various lockdowns were simulated by changing the lockdown values within +5% and -5% of the nominal. The values for lockdown variations for India are 66% and 76%, while the nominal was 71%. Similarly, for the USA, lockdown variations were 70% and 78%, while the nominal was 74%.

## 6 Discussion

MIMANSA uses multiple parameters as inputs. It is based on the current understanding of the disease. However, with the possibility of a new vaccine, herd immunity, and antibody tests, the parameters will need to be recalculated. The values of lockdown also change. Thus, in the simulations shown in the results section, we have made predictions for a limited time.

The two main parameters that control the count of the number of cases are the exposure rate and the infection rate. In section 3, equation(1) also shows that the prominent terms that could increase the virus-infected patients (VP) are the exposure rate, and the infection rate.

The exposure rate is a measure of how much a person gets exposed to a virus when in contact with a silent carrier. It is a function of multiple factors. The exposure depends on the duration of the meeting, the number of contacts, the virus activity level at the time of the contact, mask usage, social distancing, hand washing, etc.

Exposure depends on what level of precautions individuals take. People wearing a mask, keeping the prescribed social distance, and washing hands frequently will undoubtedly reduce the exposure level. The individual exposure risk goes down when one follows the recommended practices. The overall exposure levels go down for the community if the community follows the recommended practices. It is certainly possible that most people will follow these basic rules. Thus, even if one comes in contact with a carrier, the exposure rate ER (1) will be less for those who follow the rules compared to the people who do not follow the rules.

In the future, more work needs to be done on reducing the exposure rate and the infection rate. As and when more tests are conducted, we will know on how to account for immunity as an impact factor in MIMANSA.

MIMANSA is a self-driven model. As a starting point, MIMANSA needs to know the presence of silent carriers. The simulator starts with the number of silent carriers as its input. Once started, it will continue to build the virus spread network as more and more people come in contact with the initial silent carriers. In this process, it creates more and more silent carriers. These newly created silent carriers have their contacts or network. They mix with people in the other networks, and it fuels the growth of the virus. The only way to stop this cycle is to break the link in more than one way. It would involve multiple methods from isolating potential silent carriers, or reducing the exposure by social distancing, or using a mask, or washing hands frequently and touching eyes, mouth, and nose, or self-imposed isolation from others.

One of the biggest strengths of MIMANSA is that it represents the observed reality. The model is flexible and easy to modify as and when new observations are known, and new discoveries are made. At present, it uses the exposure rate and the infection rate as the main driving factor.

MIMANSA is not a COVID-19 specific model. It is a generic model that is capable of simulating any virus spread. For a different virus, one has to change the exposure rates, the applied PDFs, the incubation period, the infection rate, and perhaps some assumptions to reflect the characteristics of the new virus.

MIMANSA helps one simulate scenarios to study the impact of many different conditions. It assists public health officials in complex decision making, enables scientists in projecting the SARS-CoV-2 virus spread, and aids hospital administrators in the management of COVID-19 patients better.

## 7 Conclusion

In this paper, we have given a detailed description of how a multilevel integrated model for the spread of COVID-19 is built using the systems approach. We designed the system by keeping the real-world view in perspective at all times.

Daily in-person interactions with 22 people were split into three layers, namely family, workplace, and public places. These interactions form the basis of person-to-person virus transmission. Every day was built as a computational block connected to other layers through the silent carriers coming out of the block. Each day comprises of many layers of computation blocks.

A complex network of the virus spread was built block-by-block and layer-by-layer. The multiple layers were integrated to give a complete system view. All parameters have physical significance in the real world.

The MIMANSA simulator has four controls. They are the virus exposure control, the infection rate control, the lockdown control, and the quarantine control. Besides these, there is complete flexibility in changing other parameters, if there is empirical evidence.

In this paper, we have shown the models’ effectiveness in simulating COVID-19 However, MIMANSA is an inherently generic model to simulate the virus spread. It is flexible and can be easily adapted to other spread of viruses.

MIMANSA was trained using COVID-19 case data from the USA and India. Once trained, it has been able to track the number of patient cases within 3% error margin. However, the objective of MIMNASA is not only predicting the cases but to help make complex decisions with ease.

MIMANSA allows one to simulate various percentages of lockdown, assess the effectiveness of masks usage, contact tracing, and quarantine. One can run multiple scenarios with varying levels of each of the parameters to find out a trade-off between a high number of patients with a minimal impact on the economy vs. a low number of cases with a high impact on the economy. Our current research aims at striking a balance between the two extremes like a total shutdown vs. partial opening of businesses with due precautions.

## Data Availability

The data was obtained from the link given below.
covid19india.org
kaggle.com

https:www.covid19India.org

https://www.kaggle.com/imdevskp/corona-virus-report

## Acknowledgments

We thank Dr. Abhishek Karwa, Asst. Professor of Medicine, School of Medicine, University of California San Francisco, California, for meticulously going through multiple drafts of the paper and making valuable suggestions. His inputs have been instrumental in shaping this paper.

## 8 Appendix

